# Detection of measles virus genotype D8 in wastewater of Brussels capital region, Belgium, March 2024

**DOI:** 10.1101/2024.04.08.24305478

**Authors:** Annabel Rector, Mandy Bloemen, Bart Hoorelbeke, Marc Van Ranst, Elke Wollants

## Abstract

Following the European Centre for Disease Prevention and Control (ECDC) threat assessment brief regarding increased measles case numbers and outbreaks in Europe, we investigated sewage samples from different wastewater treatment plants in Belgium for the presence of measles virus (MV). Samples taken from Brussels North were found to contain MV, which could be characterized as genotype D8. Twelve measles cases were reported in the Brussels capital region in February/March 2024, but additional infections are likely occurring under the radar.

---

On February 16^th^ 2024, the European Centre for Disease Prevention and Control (ECDC) issued a threat assessment brief in response to the significant increase in measles case numbers and outbreaks that have been observed in 2023, globally as well as within the European Union (EU)/European Economic Area (EEA) (1). With a notification rate of 5.94 per 1 million population in 2023, Belgium was among the countries with the highest measles rate of the EU/EEA, and it’s 2 dose vaccination coverage (∼83% in 2022) is below the European average (2). Furthermore, a large outbreak of measles has been reported in the Democratic Republic of Congo (DRC) (3), which has close ties with Belgium and is connected through multiple direct flights weekly. The high probability of measles importation from regions with substantial circulation combined with the upcoming seasonal peak of the virus and sub-optimal vaccination coverage can likely result in a continued increase in measles cases in Belgium in 2024. We have recently demonstrated that wastewater surveillance can be used as an early warning for upsurges in respiratory virus circulation (4). To investigate the degree of viral circulation, we examined sewage samples from different locations in Belgium for the presence of MV.

## Sewage sampling

Wastewater samples were obtained from 3 large regional WWTP. Since December 2020, we examine wastewater samples of the plant of Aquafin Leuven on a weekly basis to track circulation levels of SARS-CoV-2 (5). This plant drains wastewater from 8 municipalities in the region of Flemish Brabant, comprising of approximately 120,000 inhabitants. In response to the ECDC threat assessment brief of February 2024, we immediately initiated measles testing on these samples. We additionally obtained wastewater samples from the Brussels South WWTP, which sanitates wastewater of the 8 Brussels municipalities, corresponding to 360,000 population equivalents, and the Brussels North WWTP, which sanitates the wastewater of the Brussels Capital Region and the Flemish Woluwe basin, covering 1.1 million population equivalents. Extracted material from two wastewater samples taken on 26/03/2024 at the WWTP of Antwerp North and South (77,000 and 216,000 population equivalents respectively) was kindly provided by Prof. Delputte of the University of Antwerp (UA)

In these WWTPs, samples of 24 hours influent wastewater were obtained with a time proportional sampler. Samples were stored at 4°C, and after transportation to the lab pretreatment and extraction of viral nucleic acids was performed as previously described (5).

## Laboratory methods for detection and genotyping of measles virus from wastewater

Molecular detection of measles virus RNA was performed by real-time RT-PCR with the TaqMan™ Fast Virus 1-Step Master Mix (Thermofisher, catalog number 4444434) using primer pair N1F (5’-CGATGACCCTGACGTTAGCA-3’) and N1R (5’-GCGAAGGTAAGGCCAGATTG-3’), with the specific probe (FAM-5’-GGCTGTTAGAGGTTGTCCAGAGTGACCAG-3’-BHQ1) described by Benschop et al. (6). Quantification was performed using a 10-fold dilution series of a 97 nt synthetic oligonucleotide covering the detection region to generate a standard curve ranging from 2.4 to 2.4 × 10^5^ copies/μl. Copies per μL extracted RNA isolated from the concentrated sewage samples correspond to copies per ml of untreated wastewater. Human faecal indicator pepper mild mottle virus (PMMoV) was quantified to normalize for differences in human waste input and changes in wastewater dilution (5).

For initial virus characterization, 295 nt in the carboxyterminal region of the nucleoprotein (N) gene of the virus were amplified and sequenced. A nested PCR approach, based on a method developed for monitoring of SARS-CoV-2 populations in wastewater, was used to allow amplification starting from the very low copy number present in the wastewater RNA extract (7). The primary RT-PCR, amplifying a 589 bp fragment, was performed with primers MVN1001-F (5′ GCTATGCCATGGGAGTAGGAGTGG 3′) and MVN1590-R (5′ GGCCTCTCGCACCTAGTCTAG 3′) (8). This was carried out using the Superscript IV One-Step RT-PCR System (Thermo Fisher Scientific,12594100) in 25 μL containing 0.5μM of each primer, 0.25 μL of SuperScript™ IV RT Mix, 12.5 μL 2X Platinum SuperFi™ RT-PCR Master Mix and 2 μL of wastewater extracted nucleic acids, with cycling conditions as follows: 2 min at 25°C, 20 min at 50°C (20 min), 2 min at 95°C followed by 30 cycles of 15 sec at 95°C, 30 sec at 55°C and 1 min at 72°C. A secondary PCR was performed with nested primers MVN1123-F (5′ CTTGTTTCAGAGATTGCAATGCAT 3′) and MVN1471-R (5′ ACCTTCGACTGTCCTGCGGATCT 3′) (8). The reaction was carried out with Q5 High-Fidelity DNA Polymerase (New England Biolabs, M0491) in 25 μL containing 0.125 μL Q5 polymerase, 4 μL Q5 Reaction Buffer, 0.5 μM primers, dNTPs (100 μM each) and 5 μl of the primary PCR product as template. Secondary PCR cycling conditions were as follows: 2 min at 95°C followed by 25 cycles of 15 sec at [95°C, 30 sec at 55°C and 1 min at 72°C. Negative controls (water) were taken along throughout the entire process. Purified amplicons were sent to the Macrogen sequencing facility in Amsterdam (The Netherlands) for Sanger sequencing with the inner forward and reverse primers. Sequences were manually corrected using Chromas 4.1, contigs were generated and primer sequences were removed in SeqMan. Analysis of the sequence with BLAST as well as with the measles typing tool on Genome Detective (https://www.genomedetective.com/app/typingtool/measles/) indicated the presence of measles virus genotype D8, which is currently circulating in Europe (9).

The WHO guidelines recommend that the sequence of the 450 nucleotides encoding the carboxyterminal 150 amino acids of the nucleoprotein should be used as the minimum amount of sequence data required for determining the MV genotype (10,11). To allow nested PCR amplification of this entire carboxyterminal region of the N gene, additional primers MVN-outF2 (5’-AAGTTCAGTGCAGGATCATAGC-3’) and MVN-outR2 (5’-TAGGTGGATGTTCTGGTCC-3’) were designed based on an alignment of GenBank sequences with the highest similarity to the initial amplicon sequence. Nested PCR was carried out as described above using primers MVN-outF2 and -R2 in the primary RT-PCR and primers MVN1001-F and MVN1590-R in the secondary PCR, followed by sequencing of the secondary PCR product.

## Measles virus in sewage water in relation to reported cases

Concentrations of MV in sewage water of the WWTP of BXL-N are shown in Table 1. Whereas no MV was detected in the wastewater of the WWTP of Leuven (21 samples investigated, taken between 28/12/2023 and 26/03/2024), Antwerp north and south (samples of 26/03/2024) and BXL-S (sample of 10/03/2024), the first sample taken in BXL-N was positive with a Cq value near the detection limit. The extremely low amount of MV genetic material in this sample did not allow sequencing for confirmation, and was therefore not reported. The result however did require the investigation of follow up samples, which were again positive and contained higher copy numbers of MV. The sample of March 8^th^ was of sufficiently high positivity to allow amplification by nested PCR and sequencing, confirming that the sample did not contain a vaccine strain. This finding, together with the rise in copy number and MV/PMMoV ratio urged us to notify public health authorities. The total number of confirmed measles cases reported in Belgium this year (up to mid-March 2024) is 36, 12 of which were in the Brussels Capital Region (data provided by Vivalis Brussels). It is not possible to reliably deduce the number of persons shedding MV from the concentrations measured in our wastewater sample. It has been estimated that investigation of sewage water would allow detection of 1 MV shedder in 5 × 10^2^ to 5 × 10^3^ persons in residential settings and schools respectively, based on the analytical detection limit of the real-time RT-PCR assay, and estimations on the amount of average measles virus urinary excretion per day and of water discharge (6). Although we have used the same analytical assay for measles quantitative detection, these numbers cannot be arbitrarily transferred to our setting since different wastewater sampling methods were used. It however strongly suggests that the MV detection in a wastewater sample covering over 1 million population equivalents originates from a substantial number of virus shedders, and indicates that the virus is probably circulating under the radar. Further research is needed to refine the relation between MV quantities in wastewater and active case numbers.

**TABLE.** Viral quantification in wastewater of Brussels north, Belgium, 2024 (n□=□5 samples)

| Sample date | MV Cq | MV copies/mL | PMMoV Cq | PMMoV copies/mL | Ratio MV/PMMV |
| --- | --- | --- | --- | --- | --- |
| 28/02/2024 | 39.65 | 2 | 21.75 | 4.6 x 10<sup>5</sup> | 4 x 10<sup>-6</sup> |
| 07/03/2024 | 38.41 | 3 | 22.68 | 2.5 x 10<sup>5</sup> | 1 x 10<sup>-5</sup> |
| 08/03/2024 | 35.63 | 22 | 21.67 | 4.8 x 10<sup>5</sup> | 5 x 10<sup>-5</sup> |
| 18/03/2024 | 38.12 | 4 | 22.56 | 2.7 x 10<sup>5</sup> | 2 x 10<sup>-5</sup> |
| 19/03/2024 | 38.75 | 3 | 22.64 | 2.5 x 10<sup>5</sup> | 1 x 10<sup>-5</sup> |

Cq: quantification cycle; MV: measles virus; PMMoV: pepper mild mottle virus

Following the ECDC recommendation to enhance public awareness about the risks of MV disease and outbreaks (1), we wanted to communicate our findings to a broader public. We therefore shared our results on X on March 14^th^ 2024 (https://twitter.com/ELwollants/status/1768227360376074655), and this was immediately picked up by national and regional media, resulting in high public exposure.

## Measles virus sequences from sewage water in comparison to reference sequences and circulating strains

Detailed phylogenetic comparison of the partial MV sequence retrieved from the BXL-N wastewater sample to reference sequences (WHO) showed highest similarity to genotype D8, which is currently circulating in Europe (Figure 1). It was most closely related to recent strains from Romania, where an outbreak has been ongoing since mid-February 2023 and a national measles epidemic was declared on December 5^th^ 2023 (1,12).

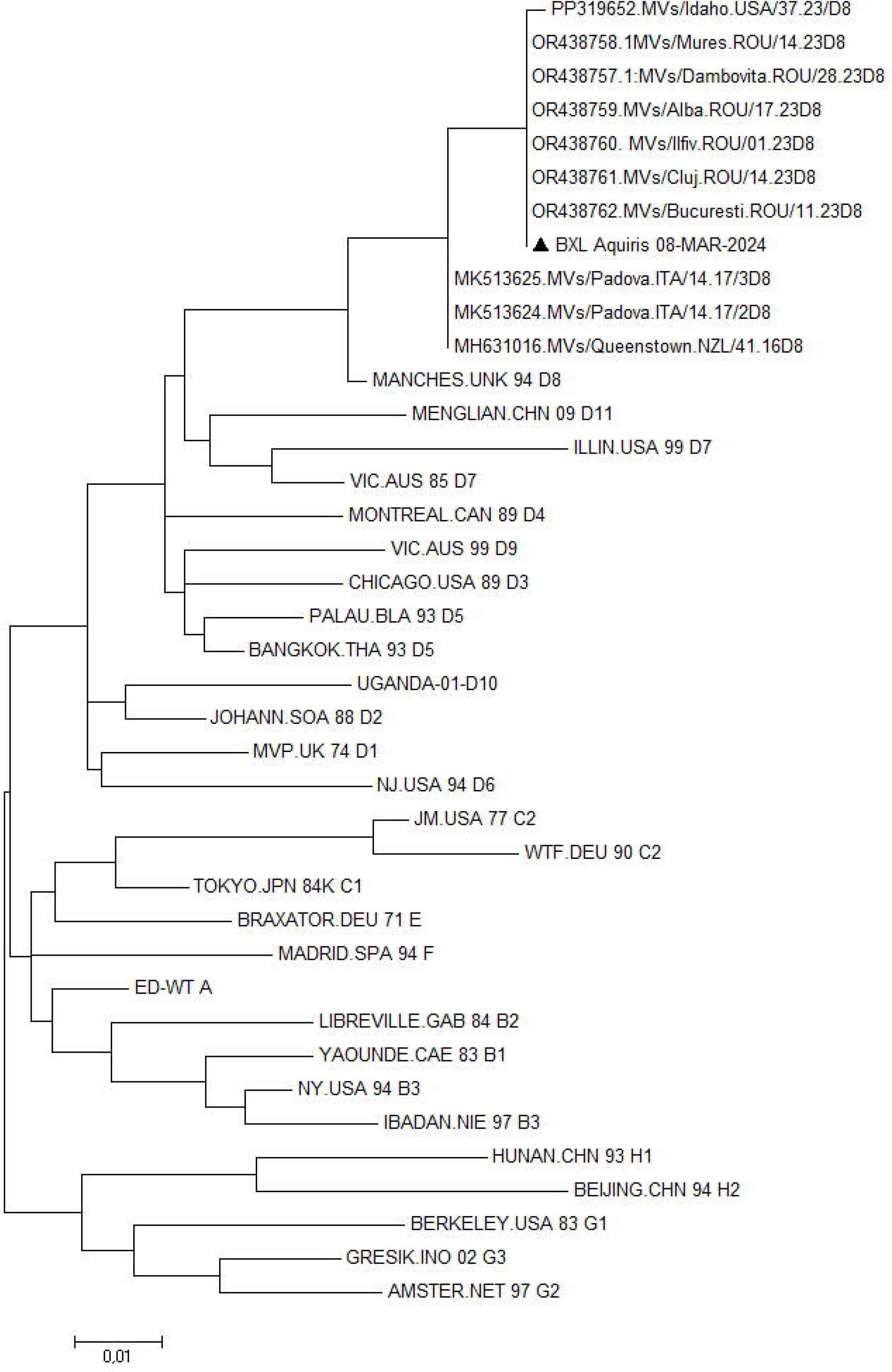
Evolutionary analyses were conducted using the 450 nucleotides coding for the C-terminal 150 amino acids of the measles virus nucleoprotein. The phylogenetic tree includes the sequence found in wastewater of Brussels north (BXL_Aquiris_08-MAR-2024), the WHO reference sequences (kindly provided by Dr. Paul Rota, CDC) and the 10 sequences with the highest similarity as retrieved by BLAST (Basic local alignment search tool) search. The evolutionary history was inferred in MEGA version 7 using the Maximum Likelihood method and and General Time Reversible model (13). The tree with the highest log likelihood is shown. Initial tree(s) for the heuristic search were obtained automatically by applying Neighbor-Join and BioNJ algorithms to a matrix of pairwise distances estimated using the Maximum Composite Likelihood (MCL) approach, and then selecting the topology with superior log likelihood value. A discrete Gamma distribution was used to model evolutionary rate differences among sites. The rate variation model allowed for some sites to be evolutionarily invariable.

In conclusion, the methodology outlined in this paper enables highly sensitive genotyping of MV from wastewater. With this method, we detected the presence of MV genotype D8 in wastewater samples of BXL-N during 3 consecutive weeks, indicating substantial viral circulation in this region. We believe that wastewater testing for measles can offer a valuable surveillance tool to trace hot spots of virus circulation.

## Data Availability

All data produced in the present study are available upon reasonable request to the authors

